# Photobiomodulation for postoperative pain relief in endodontic surgery: a randomized controlled study protocol

**DOI:** 10.1101/2023.10.18.23297226

**Authors:** Rolf Wilhem Consolandich Cirisola, Luis Eduardo Pascuali Moya, María Victoria García Olazabal, Daniela Anat Amzallag Wagmann, Guendalina Palermo Suarez, Carolina Wince, Laura Hermida Bruno, Daniel Rodriguez Salaberry, Maria Cristina Chavantes, Priscila Larcher Longo, Lara Jansiski Motta, Sandra Kalil Bussadori, Cinthya Cosme Gutierrez Duran, Kristianne Porta Santos Fernandes, Raquel Agnelli Mesquita Ferrari, Anna Carolina Ratto Tempestini Horliana

**Affiliations:** Postgraduate Program in Biophotonics-Medicine, Universidade Nove de Julho (UNINOVE), São Paulo, Brazil; Universidad Católica del Uruguay (UCU), Montevideo, Uruguay; Postgraduate Program in Aging Science, Universidade São Judas Tadeu (USJT), São Paulo, Brazil; Postgraduate Program in Reabilitation Sciences, Universidade Nove de Julho (UNINOVE), São Paulo, Brazil

**Keywords:** photobiomodulation, endodontic surgery, pain, inflammation

## Abstract

Photobiomodulation (PBM) has shown favorable results in the postoperative period of endodontic surgery, however, up to now, the level of evidence in this procedure is low. The objective of this study will be to evaluate if photobiomodulation (PBM) can reduce postoperative pain in patients who will undergo endodontic surgery. For this randomized, controlled, and double-blind clinical study, 34 patients without comorbidities who need endodontic surgery in the upper jaw (15 to 25 teeth) will be recruited. They will be randomly divided into an experimental group (n = 17) photobiomodulation (808nm, 100 mW, and 4J/cm^2^ with 5 points per vestibular). Applications will be made in the immediate postoperative period and 24 hours after surgery. Control group (n = 17) PBM simulation will be performed in the same way as in the experimental group. In this group, the required analgesia will be administered within the standard with ibuprofen. Both groups will perform the necessary conventional procedures considered the gold standard in the literature. Both the patient and the evaluator will be blinded to the intervention performed. The primary outcome variable of the study will be postoperative pain, which will be assessed using the visual analogue scale at all postoperative control visits (baseline, 24 hours and 7 days). As for the secondary outcome variables, the amount of systemic medication received according to the patient’s need (will be provided by the investigator). Radiographic images will be obtained after 1 and 3 months for evaluation of the repair (dimensions of the lesion, radiopacity). These radiographs will be taken digitally with the positioners implemented. Edema, ecchymosis, and evaluation of soft tissues in the anterior portion of the intra and extraoral maxilla will also be evaluated. In addition, the temperature with a digital thermometer. These parameters will be evaluated 24 hours and 7 days after the intervention. The intervention and the X-rays will be taken in the 1^st^ and ^3rd^ month respectively. Once all the data have been collected, their normality will be tested, and the one-way ANOVA test and the complementary Tukey test will be carried out. Data will be presented as mean ± standard deviation (SD) and the accepted p-value will be <0.05.

## Introduction

The objective of endodontic surgery is to surgically access the defect, remove the lesion and affected tissues, perform the resection of the fragment at the level of the involved root apex, location of the involved canal(s), retro conformation, three-dimensional cleaning and sealing of all the ends of the root canal (retro filling) with a biomaterial. (Forbes et al., 2000). The success rate in endodontic surgery has improved significantly from 60% to 90% (Von Arx et al., 1999; Zuolo et al., 2000). The postoperative period can be painful depending on the magnitude of the surgical trauma, the presence of microorganisms, and non-compliance with postoperative recommendations. In general, there is little literature related to this type of surgery and the possible results come mainly from oral or periodontal surgery and not from endodontic surgery (Taschieri, 2021).

NSAIDs are the most prescribed drugs to prevent pain (Nekoofar et al., 2003), with ibuprofen being the most researched drug. Evidence has shown that NSAIDs should be avoided in patients at high risk of cardiovascular disease. If used, it should be done in shorter doses and durations, with the goal of achieving maximum effectiveness (Bally et al., 2017). If used, it should be done in shorter doses and periods of time, with the aim of achieving maximum effectiveness. In recent years, photobiomodulation has been used successfully in oral surgery to reduce edema and postoperative pain, however, there are few studies that have tested it in the postoperative period of endodontic surgery.

Photobiomodulation has been widely used in oral surgeries with promising results, however, in the postoperative period of endodontic surgeries it has not been widely studied. Low power laser application has been used for several decades to reduce pain, and inflammation and improve wound healing conditions (Karu 2014, Hamblin 2017) and it is well-established in the dental clinic due to its anti-inflammatory and regenerative effects. (Okamoto et al., 1993; Roberts-Harry et al., 1992). LLLT is considered an adjunct to relieve postoperative pain from the procedure (Turhani et al., 2006). There are few studies that have studied the effect of photobiomodulation on periapical tissue repair after endodontic surgery (Metin et al., 2018; Payer et al., 2005; Kreisler et al., 2004).

Regarding the parameters used for irradiation, the radiant exposure ranged between 3 and 15 J/cm^2^. In one of the studies (Metin et al., 2018) 3.87 J/cm^2^ was used, while in the other study (Payer et al., 2005) 3-4 J/cm^2^ was used. In the study by Kreisler et al., 2004, only the amount of energy per point (7.5 J) and the fiber diameter (600 μm) were reported. Achieving a radiant exposure of 15 J/cm^2^. Despite the good results, molars were included in the sample. Payer’s study did not perform well for the photobiomodulation group, however, the wavelength used was 680 nm, which is not ideal considering the depth light needs to penetrate. Metin’s study, for its part, used infrared (810nm) with 3.87 J/cm2 of radiant exposure and included only anterior teeth in the sample, so it was used as a reference for our parameters. We use them as a reference because they are common in clinical practice and within the parameters reported in the literature in studies with the least amount of bias and with good results (Metin et al., 2018).

Taking all these factors into account, it is suggested that photobiomodulation may be an effective alternative to reduce the use of NSAIDs in pain control after endodontic surgery, thus eliminating the adverse effects of these drugs (Nabi et al.,) and may be tested in the future in systemically compromised patients. Therefore, the objective of this study will be to evaluate if photobiomodulation can reduce postoperative pain after endodontic surgery compared to conventional treatment (ibuprofen) using the visual analog scale (VAS) at the beginning of the study, 24 hours and 7 days after surgery.

## Material and methods

This randomized, controlled, double-blind superiority clinical trial meets the criteria for designing a clinical trial in accordance with the SPIRIT Statement. It was accepted by the Research Ethics Committee (CEP) of the Catholic University of Uruguay, process: 220914.

Patients who consult at the UCU Clinic and require an endodontic surgery as treatment will be referred to the Project. After the verbal and written explanation of the study, the participants who agree to participate will sign the Informed Consent Form.

The treatments will be carried out in the surgical unit of the Clínica Universitaria de la Salud of the Universidad Católica del Uruguay in Montevideo city, Uruguay, from 30^th^ October 2023 to August 2024. Any complications or changes will be reported and clarified to the CEP and reported in publications. Personal information of participants will be collected, shared, and safeguarded for confidentiality throughout the trial

### Sample description

Participants of both sexes who previously consulted at the Clínica Universitaria de la Salud, with a diagnosis of periodontitis with an apical lesion less than 10mm with or without a fistula, diagnosed clinically and radiographically, in the upper maxillary region (from 15 to 25). They will be invited by the principal researcher, who will obtain informed consent.

All patients will receive endodontic surgery as the indicated therapeutic means with no difference between the procedures except for the application of light.

The PBM will be performed after surgery and 24 hours and 7 days after surgery.

### Inclusion/exclusion criteria

Participants will include:

- Patients with periapical lesions who have already undergone endodontic treatment (lesions smaller than 10mm in their greatest diameter - Metin et al., 2018, single and chronic lesions)
- Patients with no comorbidities,
- Age from 18 to 70 years,
- Both genders,
- Healthy permanent teeth with good hygiene.

### Participants will be excluded

- Who are taking drugs that affect bone metabolism and the inflammatory process (for example: corticosteroids, bisphosphonates),
- Smokers, pregnant or lactating women,
- Who used anti-inflammatory drugs in the last 3 months before surgery.
- Who for any reason interrupted the evolution of the treatment for not attending joint appointments.
- Patients who do not follow the guidelines or have an injury in the acute phase (pain, edema, exudate)

Those patients who may present some complication during the research period will be assisted at the UCU Clinic or, if outside of hours, at the Mobile Coronary Unit within the framework of the agreement between the UCU and the Mobile Coronary Unit (UCM). All patients will be informed that the adverse effects that may occur will be inherent to the surgical procedure and their resolution will be carried out according to the usual protocol for these cases.

If during the scheduled consultations, the patient moves to another city or simply does not want to participate in the survey, they will be automatically disconnected, without any commitment or damage to it.

Data from all patients randomized to the survey will be included in the statistical analysis, described and discussed, as well as potential adverse effects. We will use intention-to-treat analysis. Participants will receive assistance from the researchers for all problems arising from the research.

### Sample’s size calculation

The total sample size will be 34 patients per group. This value was calculated to give a power of 95% (α = 0.05) and an effect size of 0.6421598. To determine the number of patients in each group, a sample calculation was performed based on the variability of the results of 1 article that evaluated the pain outcome, measured in millimeters (mm) with the visual analog scale. The same time interval as the study (24 h) was considered. In one group, PBM was used and the mean pain in millimeters was 1.91 ± 1.76 and the other group did not use PBM, and the mean pain was 3.14 ± 2.04. Identical situation to the primary outcome measure used in this study. Using the two-tailed t-test method, the required sample will be 34 individuals, 17 per group. Calculations were made using a significance level of 0.05 (implying a type I error of 5% and leading to an analysis with a 95% confidence interval) and an absolute error of 5%. The flow diagram of the study, according to the SPIRIT recommendations, is shown in Figure 1

**Figure 1.**
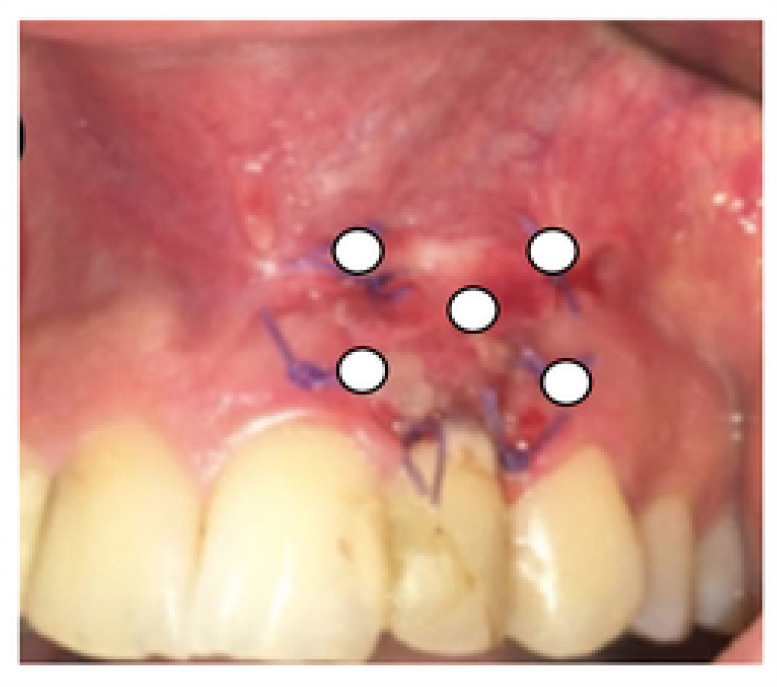

### Calibration and examiner training

One examiner will be trained to assess visual analog scale, temperature assessment in order to maximize reproducibility of assessments.

### Randomization

The treatment carried out immediately after surgery will be randomized and may be photobiomodulation or photobiomodulation simulation. A program available on the internet and a random sequence generator (https://www.sealedenvelope.com/) will be used and the option of randomization by blocks of 2 treatments will be selected. The opaque envelopes will be identified with sequential numbers and the information of the corresponding experimental group will be inserted inside, and sealed. The generation of the random sequence and the preparation of the envelopes will be carried out by a person not directly involved in the study. Immediately after finishing the suture, the investigator in charge of applying the PBM will remove and open 1 envelope (without changing the numerical sequence of the other envelopes) and perform the indicated procedure or its simulation. Only this researcher will know the nature of the treatments.

### Group composition

#### G1-SHAM group

Conventional treatment + PBM simulation (n = 17 patients): All participants will undergo the same conventional surgical procedure. Patients will receive the PBM simulation and will be treated identically to the G2 group as shown in Figure 2. The person responsible for applying the PBM will simulate the radiation by placing the devices in the same places described for the PBM group, however, the equipment will remain turned off. Thus, to the participant does not identify the group to which he belongs, the device activation sound (beep) will be recorded, and it will turn on at the time of application.

**Figure 2.**
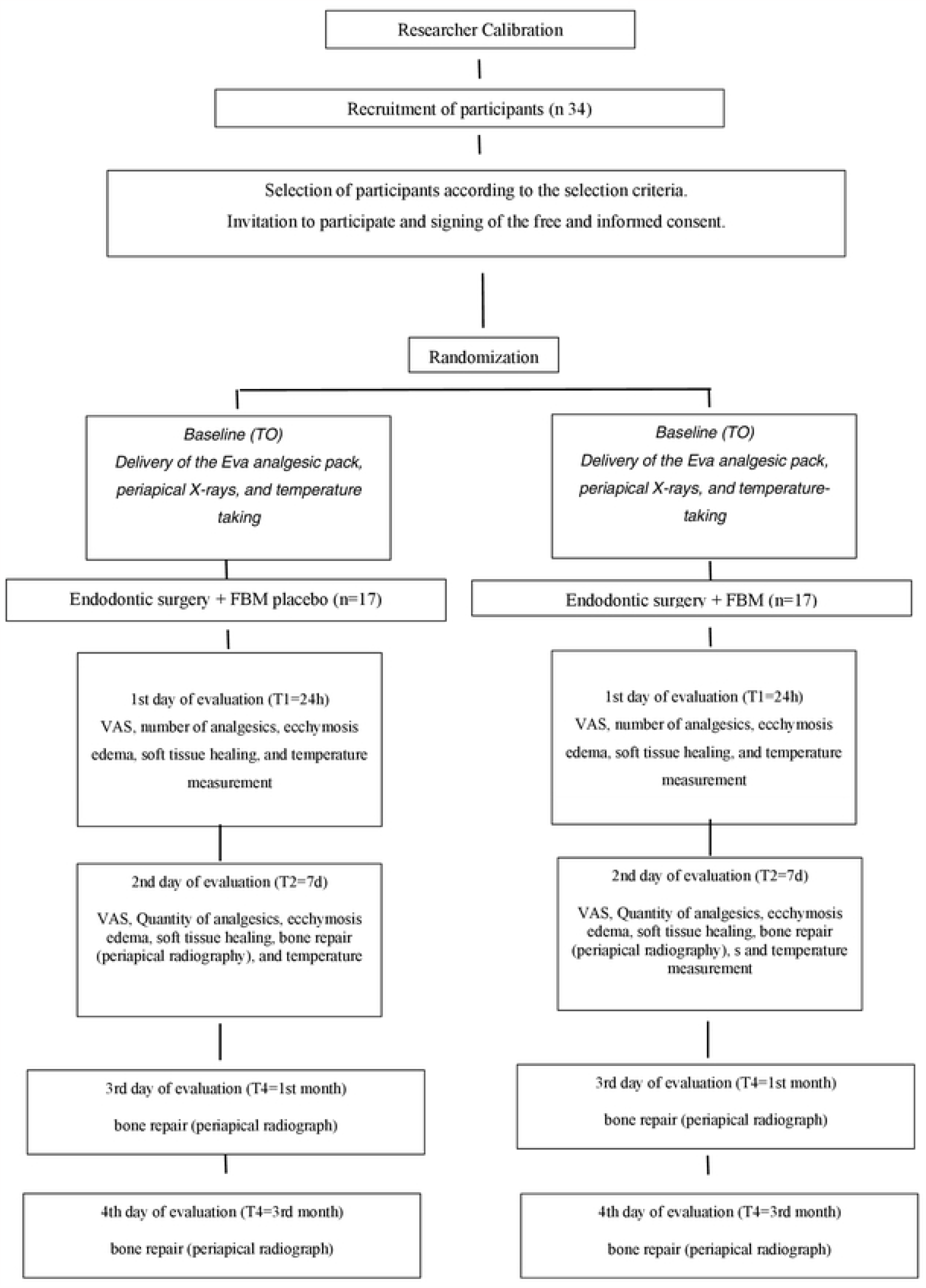
Regions of laser application on the vestibular face (Source: adapted from Metin et al 2018)

#### G2-Intervention group

Conventional treatment with placebo ibuprofen + PBM (n = 17) All participants will undergo the same surgical procedure. Patients will receive PBM (Table 1) and will be treated identically to the G2 group.

**Table 1:** Dosimetric parameters (modified from Metin et al., 2018 and Payer et al., 2005.

| Parameters | Values/treatment |
| --- | --- |
| Wave compression [nm] | 808 |
| operating mode | continuous |
| Radiant power [mW] | 100 |
| Irradiance [mW/cm<sup>2</sup>] | 200 |
| Beam area [cm<sup>2</sup>] | 0.5 |
| Exposure time [s] | twenty |
| Radiant exposure [J/cm<sup>2</sup>] | 4 |
| Radiant energy [J] | 2J per point |
| Total energy [J] | 10J |
| Number of irradiated points | 5 points for vestibular. Irradiate 1 point in the center of the lesion and equidistantly around it, perform 4 irradiations in a quadrangular fashion. With a distance of 1 cm between them. |
| application technique | In contact, 90 degrees to the surface |
| Number of sessions and frequency | Postoperative, 24 hours and 7 days |

The irradiated region will be on the lesion at 4 equidistant points on the vertex of a flat square 1 cm away. A dot will be irradiated in the middle of the square (Figure 3). Placebo ibuprofen will be manipulated.

**Figure 3.**
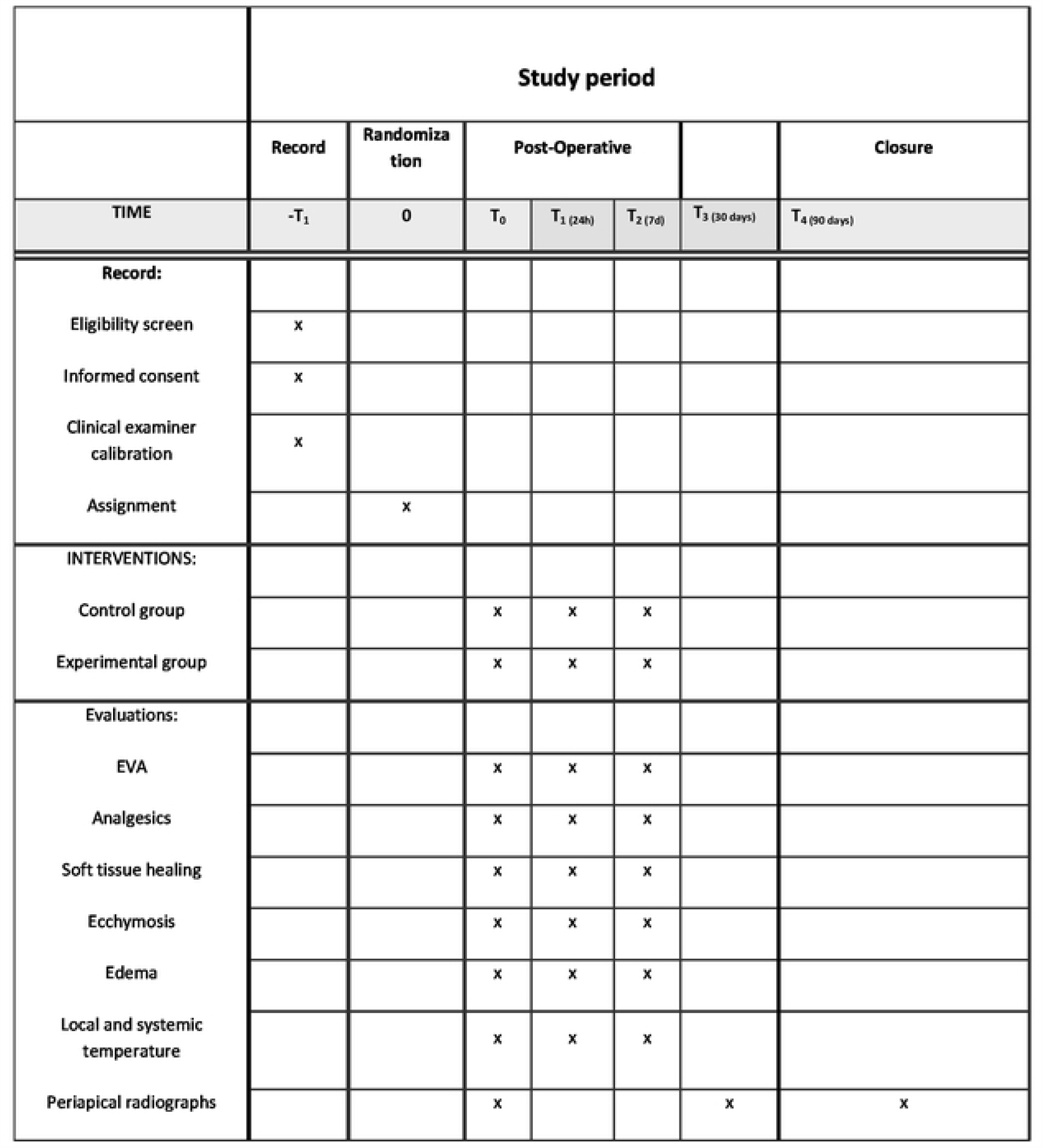

Dosimetric parameters and the number of PBM applications are described in Table 1.

### Study Outcomes

The primary outcome of the study will be:

- **Pain** (comparison of the laser group with the conventional treatment group with ibuprofen) during the immediate postoperative period of endodontic surgery (at baseline, 24 hours and 7 days) using the Visual Analogue Scale (VAS) measured in millimeters (Metin et al.., 2018, Sampaio-Filho et al., 2018).

### The secondary results of the study will be

- **Quantity of painkillers ingested in the period:** the quantity of painkillers ingested in the 24-hour and 7-day periods will be counted. The analgesic used will be only paracetamol that will be administered to the patient, but it is advisable to take it only in case of pain (Sampaio-Filho et al., 2018). A procedure will be carried out to monitor the adherence of the participants (for example, each patient will be asked to bring the pain reliever pack to the appointment to see how it is used).
- **Edema:** a scale will be used to quantify the amount of edema as recommended by some authors (Metin et al., 2018). The scale is made up of scores from 0 to 3, where: 0 = no edema, 1 = intraoral edema, 2 = extraoral edema, and 3 = diffuse edema (Metin et al., 2018). This outcome will be assessed at baseline and within 24-hour and 7-day periods (Metin et al., 2018).
- **Ecchymosis** - bleeding in the subcutaneous tissue, with a diameter greater than 1 cm, which is caused by the rupture of one or more blood capillaries, and one of the causes is surgical trauma. Ecchymosis: 0 = no color change, 1 = spot smaller than 4 cm in diameter, 2 = spot 4-10 cm in diameter, 3 = spot larger than 10 cm in diameter (Metin et al., 2018). This outcome will be assessed at baseline and within 24-hour and 7-day periods (Metin et al., 2018).
- **Soft tissue healing:** Score 1: no opening at the incision line, no drainage (pus or exudate), no inflammation, no pain. Score 2: no opening at the incision line, no drainage, mild swelling, mild pain. Score 3: no opening at the incision line, active drainage, advanced inflammation, moderate to advanced pain. Score 4: opening at the incision line, active drainage, advanced inflammation, ongoing pain. This outcome will be assessed at baseline and within 24-hour and 7-day periods (Metin et al., 2018).
- **Bone consolidation:** periapical radiography will evaluate the changes in the area of the defect (volume and bone density). Periapical radiographs will always be performed with the same equipment using the parallelism technique in the immediate preoperative period (baseline) so that they can be compared with those performed at 7 days, 1 month, and 3 months. The area of the defect will be measured by multiplying the longest mesiodistal and super inferior diameters on the radiographs. In all radiographs, the longest diameter of the lesion was measured, and the periapical index was evaluated. The periapical index was recorded according to the following parameters: 0 = no lesion, 1 = periapical radiolucency with a diameter of 0.5-1 mm, 2 = periapical radiolucency with a diameter of 1.1-2 mm, 3 = radiolucency periapical with a diameter of 2.1-4 mm, 4 = periapical radiolucency with a diameter of 4.1-8 mm and 5 = periapical radiolucency greater than 8.1 mm in diameter. This outcome will be assessed at baseline and within 24-hour and 7-day periods (Metin et al., 2018).

### Local temperature measurement

Measurement of temperature l (local to surgery) and systemic (glabella), (comparison of the laser group with the group of conventional treatment with ibuprofen.

In the immediate postoperative period of endodontic surgery (at the beginning, 24 hours and 7 days)

### Analysis of results

Initial descriptive analyzes will be carried out considering all the variables measured in the study, both quantitative (mean and standard deviation) and qualitative (frequencies and percentages). Subsequently, normality analyzes will be carried out to determine the appropriate statistical tests for each data set (parametric or non-parametric) and the statistical tests will be applied for each specific analysis. In all tests, the significance level of 5% probability or the corresponding p-value will be adopted. All analyses will be performed using the statistical program SPSS for Windows, version.

### Monitoring

Investigators do not plan interim analysis because it is not expected serious adverse events. However adverse events (major and minor) will be collected.

## Data Availability

All relevant data from this study will be made available upon study completion

